# Divergence from Diffusion Tensor: a discriminant diffusion marker from single shell MRI

**DOI:** 10.64898/2026.10.26.26366463

**Authors:** Santiago Aja-Fernández, Antonio Tristán-Vega

**Affiliations:** LPI-BIVa, Universidad de Valladolid, Valladolid, Spain; Valladolid Health Research Institute (IBioVALL), 47010 Valladolid, Spain

**Keywords:** Diffusion MRI, fractional anisotropy, diffusion tensor, white matter

## Abstract

**Purpose:** The Divergence from the Diffusion Tensor (DDT) is a new diffusion MRI metric that quantifies how the measured diffusion signal separates from the tensor assumption. It is marker for tissue heterogeneity that is estimated from single-shell data and can be used in brain studies.

**Methods:** There exist other techniques that allow the calculation of how the measured diffusion diverges from the Gaussian and tensor assumptions. However, the calculation requires large data-sets with different b-values, which makes them incompatible which standard diffusion tensor acquisitions. By imposing an initial assumption, an alternative formulation based on one single shell (ie, single b-value) is reported.

**Results:** Publicly available databases are used to test the behavior of DDT against other anisotropy measures. It shows the ability to discriminate different non-Gaussian diffusion scenarios with very low intra-subject variabilities.

**Conclusions:** DDT can extract relevant information from the white matter, without any additional demands on data acquisition and totally compatible with standard diffusion MRI analysis pipelines. This makes DDT a good candidate to be used as an imaging marker for brain studies.

## 1 INTRODUCTION

Diffusion Magnetic Resonance Imaging (dMRI) is an imaging modality employed to assess diverse in vivo physiological and pathophysiological conditions of the human body in clinical studies. It has been widely used in the study of the brain and neurological disorders [1, 2, 3] where it allows the characterization of the diffusivity of water molecules within the tissue, providing information about the microscopic configuration and structural connectivity of the brain, especially inside the white matter (WM). There is also a current trend to use the information provided by dMRI in individual studies by quantifying the deviation of a specific brain from the norm [4, 5].

The most relevant feature of dMRI is its ability to measure directional variance in the diffusion profile, i.e., anisotropy. Different mathematical representations are used to describe the relationship between the acquired diffusion signal and the properties of the tissue under study. The most common way to estimate the anisotropy is via the diffusion tensor (DT) [6, 7]. To carry out clinical studies, the information provided by the DT is commonly translated into some scalar measures that describe different features of diffusion within every voxel. That way, metrics like fractional anisotropy (FA) were defined and widely employed to characterize damaged tissues in multiple neurological and psychiatric disorders [8, 9, 10, 11]. However, from the early stages of DT imaging (DTI), it was clear that the monomodal Gaussian assumption used by the DT approach oversimplifies the diffusion process and hence presents important limitations. Different techniques were proposed to overcome the limitations of DTI, which usually includes the acquisition of larger amounts of diffusion data. Some examples of these techniques can be seen at [12, 13].

The final purpose of these techniques is to better describe the diffusion process in tissues, by alleviating the constraints imposed by the DT assumption. Diffusion metrics based micro-structural properties like the axonal size, density, or dispersion [14, 15, 16], were this proposed, as well as parametric representations of the non-Gaussian diffusion, like the Ensemble Average Propagator (EAP) [17, 18, 19].

In addition, some of these techniques also allow the calculation of metrics that quantifies how the measured diffusion diverges from the Gaussian assumption. It is the case of Diffusion Kurtosis Imaging (DKI) [20] that includes an excess kurtosis term to capture the deviation from the Gaussian distribution. The Mean Kurtosis (MK) can be seen as a scalar measure of how the diffusion separates from a pure tensor model. It has proved to be also a good marker for tissue heterogeneity that can be used in several brain pathologies such as ischemia and infarction quantification [21], traumatic brain injury [22] or the study of neurodegenerative diseases [23].

In the EAP framework, in [18] authors proposed the non-Gaussianity (NG), a description of the (dis)similarity between the measured propagator and its Gaussian counterpart. It quantifies the consequence of micro-structural obstacles on the diffusion’s Gaussianity. It has proved its clinical applicability in the quantification of gliomas and quantification of demyelination [24]. Similar formulation for NG was also proposed in the MiSFIT approach in [25].

It seems clear that the quantification of how the acquired data separates from the initial assumptions can provide very relevant information about the diffusion process, especially in pathological cases. However, the measures in the literature, such as MK or NG, need at least 2 shells to estimate the non-Gaussian component.

In the present work we propose a new measure that we call *Divergence from the Diffusion Tensor* (DDT), which aims to quantify how the measured diffusion separates from the tensor assumption. For this purpose, we will use data from a single shell, with acquisitions fully compatible with DTI. In the present work, two different implementations are proposed.

## 2 THEORY

### 2.1 The Diffusion signal

The EAP, *P* (**R**), is the three dimensional Probability Density Function (PDF) of the water molecules inside a voxel moving an effective distance **R** in an effective time *τ*. It is related to the normalized magnitude image provided by the MRI scanner, *E*(**q**), by the Fourier transform [26]:

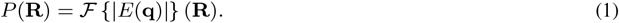

The inference of exact information on the **R**–space would require the sampling of the whole **q**–space to use the Fourier relationship between both spaces. In order to obtain a closed-form analytical solution from a reduced number of acquired images, a model for the diffusion behavior must be adopted. The most common techniques rely on the assumption of a Gaussian diffusion profile and a steady state regime of the diffusion process that yields to the well-known Diffusion Tensor (DT) approach:

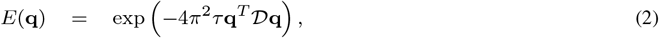

which is the well–known Stejskal–Tanner equation [27]. The diffusion tensor *D* is the anisotropic covariance matrix of the Gaussian PDF *P* (**R**), and therefore is a symmetric, positive–definite matrix, with positive eigenvalues and orthonormal eigenvectors. Eq. (2 The Diffusion signal) is usually re–written in terms of the *b*–value:

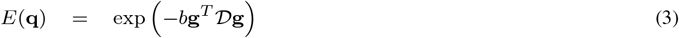

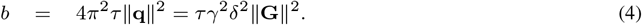

where 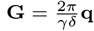 and **g** = **G***/* ∥**G**∥ = **q***/* ∥**q**∥ is the gradient direction. Alternatively, a more general exponential expression for *E*(**q**) can be used [28]:

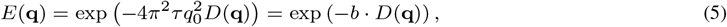

where the positive function *D*(**q**) = *D*(*q*_0_, *θ, φ*) *>* 0 is the Apparent Diffusion Coefficient (ADC) and *q*_0_ = **q**, and *θ, φ* are the angular coordinates in the spherical system. According to [29], in the mammalian brain, this mono-exponential model is predominant for values of b up to 2,000 s/mm^2^ and it can be extended to higher values (up to 3,000 s/mm^2^) if appropriate multi-compartment models of diffusion are used.

The estimation of a given magnitude is always a trade-off between the available data and the complexity of the model. In this case, we consider a single shell acquisition, since the amount of data is reduced, we are forced to assume a restricted diffusion model consistent with single-shell acquisitions: the ADC will be roughly independent from the radial direction within the range of b-values probed, so that *D*(**q**) = *D*(**u**). This way, Eq. (5 The Diffusion signal) becomes:

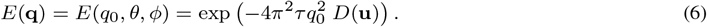

This is precisely the same assumption used by the authors in [30, 31] to estimate apparent versions of measures like the return-to-origin probability (RTOP), or the propagator anisotropy (PA), and it is the assumption used in this work.

## 3 METHODS

### 3.1 Divergence from diffusion tensor

Our intention is to quantify how the acquired diffusion signal diverges from the tensor approach, see an example in Fig. 1 Divergence from diffusion tensor. A complete measurement of this divergence would imply using the original acquired signal *E*(**q**) (without assuming any model) and the signal reconstructed after using the diffusion tensor in Eq. (3 The Diffusion signal). However, to do so, information of more than one shell is needed. In order to use just one shell, we need to impose a prior assumption to the behavior of the signal. For the sake of simplicity, we will assume that *E*(**q**) shows an exponential decay as shown in Eq. (6 The Diffusion signal), and that the diffusion signal *D*(**q**) is independent from the radial direction, *D*(**q**) = *D*(**u**). We also define the tensor representation of the acquired diffusivity, *D*_*G*_(**u**) as

**FIGURE 1.**
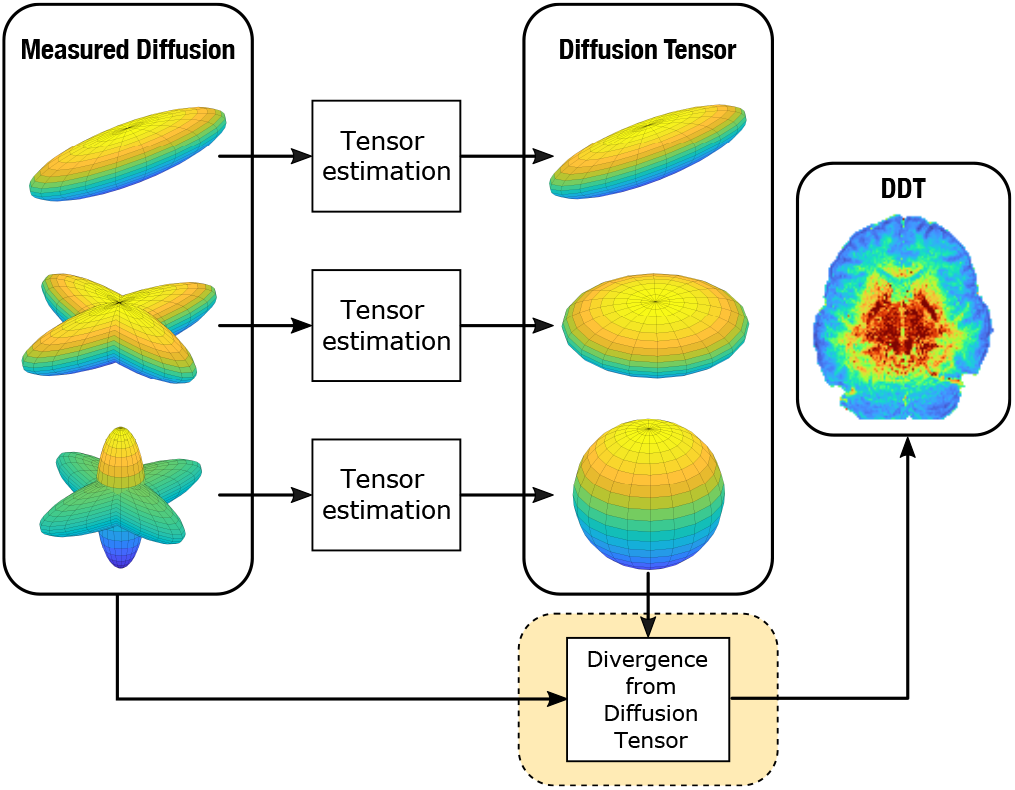
Scheme of the calculation of the DDT.

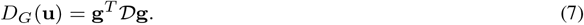

We define the *Divergence from the Diffusion tensor* (DDT) as the sine of the angle between the original signal *D*(**u**) and the tensor representation *D*_*G*_(**u**):

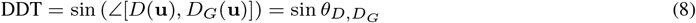

The sine is calculated from the cosine, which measures the *similarity* between two signals. It is defined as the projection of signal *D*(**u**) over *D*_*G*_(**u**) using the inner product [32, 18, 31]:

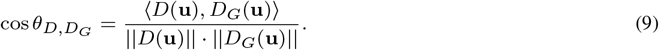

This way, the inner product of both signals is calculated as

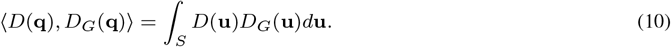

where *S* denotes the surface of a sphere of radius one. The norm of *D*(**u**) and *D*_*G*_(**u**):

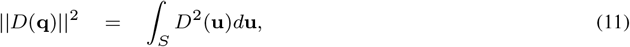

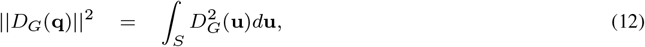

The DDT is defined straightforwardly as:

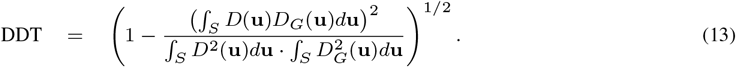

### 3.2 Practical implementation

We can represent the signal *D*(**u**) on a a Spherical Harmonics (SH) base or order *L* as

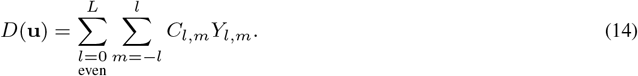

We can define *D*_*G*_(**u**) in this base by using only 2 coefficients :

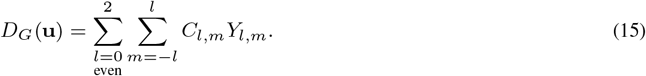

This way, we can write the integral in eq. (13 Divergence from diffusion tensor) as

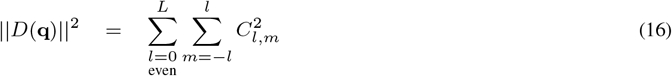

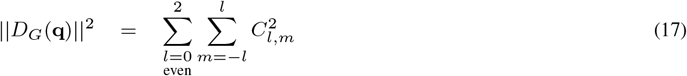

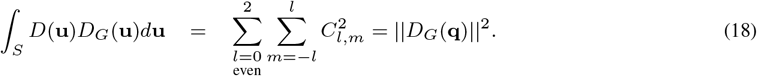

Then, eq. (13 Divergence from diffusion tensor) can be written as

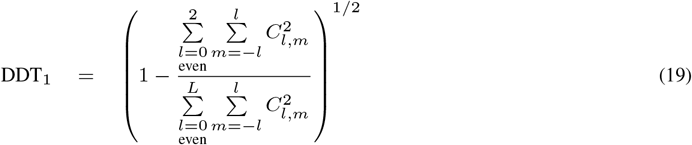

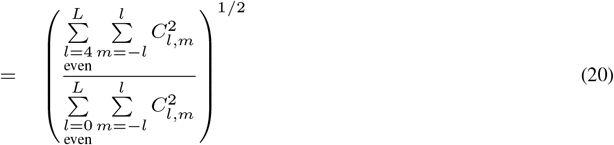

Note that the numerical implementation in eq. (20 Practical implementation) depends on the higher coefficients of the SH base (*l* ≥ 4). This way, the resulting metric can be more sensitive to noise and outliers. In what follows, we propose an alternative implementation that, albeit being optimal, could improve the robustness of the measure. We consider that the zeroth-order coefficient of a spherical harmonics (SH) expansion of signal *H*(**u**), *C*_0,0_ *{H*(**u**)}, defined as:

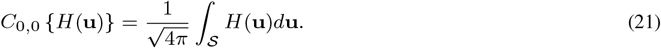

This way, we can rewrite Eq. (13 Divergence from diffusion tensor) as a function of this coefficient for different signals:

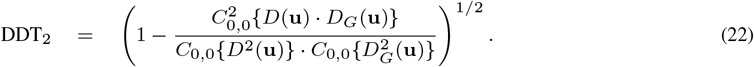

Note that, for the sake of distinction, we have denoted DDT_1_ to the first implementation and DDT_2_ to the latter.

### 3.3 Data sets

In order to test the proposed measures, three different data sets were used:

- **Human Connectome Project (HCP) database**^1^: specifically volume MGH1007, acquired on a Siemens 3T Connectom scanner with 4 different shells at *b* = [1000, 3000, 5000, 10000] s/mm^2^, with [64, 64, 128, 256] gradient directions each, in-plane resolution 1.5 mm and slice thickness was 1.5 mm.
- **HCP WU-Minn test-retest database:** [33]: In total, thirty-seven subjects were used after removing seven subjects from the database due to differences in acquisition protocol between test and retest scans (excluded cases: 135528, 137128, 151526, 169343, 179548, 192439, 601127, 660951). The subjects were scanned with a Siemens 3T Connectome Skyra scanner (Siemens, Erlangen, Germany) with a maximum gradient strength at 100 mT/m. The acquisition protocol covers three shells at *b* = [1000, 2000, 3000] s/mm^2^, 90 diffusion sensitizing gradients per each shell, and 18 repeated acquisitions at *b* = 0. Other acquisition parameters are: pulse separation time Δ = 43 ms, diffusion gradients length *δ* = 10.6 ms, voxel resolution 1.25 *×* 1.25 *×* 1.25 mm^3^.
- The **MICRA** (Micro-structural Image Compilation with Repeated Acquisitions) data set [34] contains five repeated sets of MSS diffusion-weighted MRI acquisitions for each of six healthy human participants. The images were acquired within a two-week period, approximately at the same time for each participant —avoiding potential diurnal effects— on an ultra-strong gradient 3T Connectome MRI scanner using a single-shot spin echo EPI with TR*/*TE = 3000*/*59 ms, six different shells at *b* = [200, 500, 1200, 2400, 4000, 6000] s/mm^2^, with [20, 20, 30, 61, 61, 61] gradient directions respectively, in-plane resolution 2 *×* 2 mm^2^, slice thickness 2 mm and Δ*/δ* = 24*/*7 ms. The data were preprocessed by the using the preprocessing pipeline, consisting on a removal of Gibbs ringing artifacts [35] (with MRtrix3 [36]), susceptibility-induced distortion corrections using the FSL topup tool (Analysis Group, FMRIB, Oxford, UK.; https://fsl.fmrib.ox.ac.uk/fsl/fslwiki) [37, 38] and B1 field inhomogeneity correction [39] (with MRtrix3).

### 3.4 Setting-up of the experiments

The calculation of DDT rely on the expansion of spherical functions at a given shell in the basis of SH. The fitting uses a Laplace-Beltrami penalty *λ* = 10^−6^. DDT is compared with other multishell measures of non-Gaussian diffusion, specifically NG and MK. NG is calculated with MAPL [17] using the DIPY library under Python 3.6.4. MK is calculated using the Kurtosis tensor estimated with MRtrix3 [36].

## 4 RESULTS

### 4.1 Visual Assessment

A preliminary visual assessment of the different metrics was performed using three slices from the HCP volume MGH1007. DDT was calculated using a single shell at b=1000 s/mm^2^ and b=3000 s/mm^2^. For the sake of comparison, we have also calculated NG (using shells at *b* = [1000, 3000, 5000] s/mm^2^) and MK (at *b* = [1000, 3000] s/mm^2^). Results are shown in Fig. 2 Visual Assessment. Since their ranges of variation are different from each other and each moment, all the slices shown have been min-max normalized.

**FIGURE 2.**
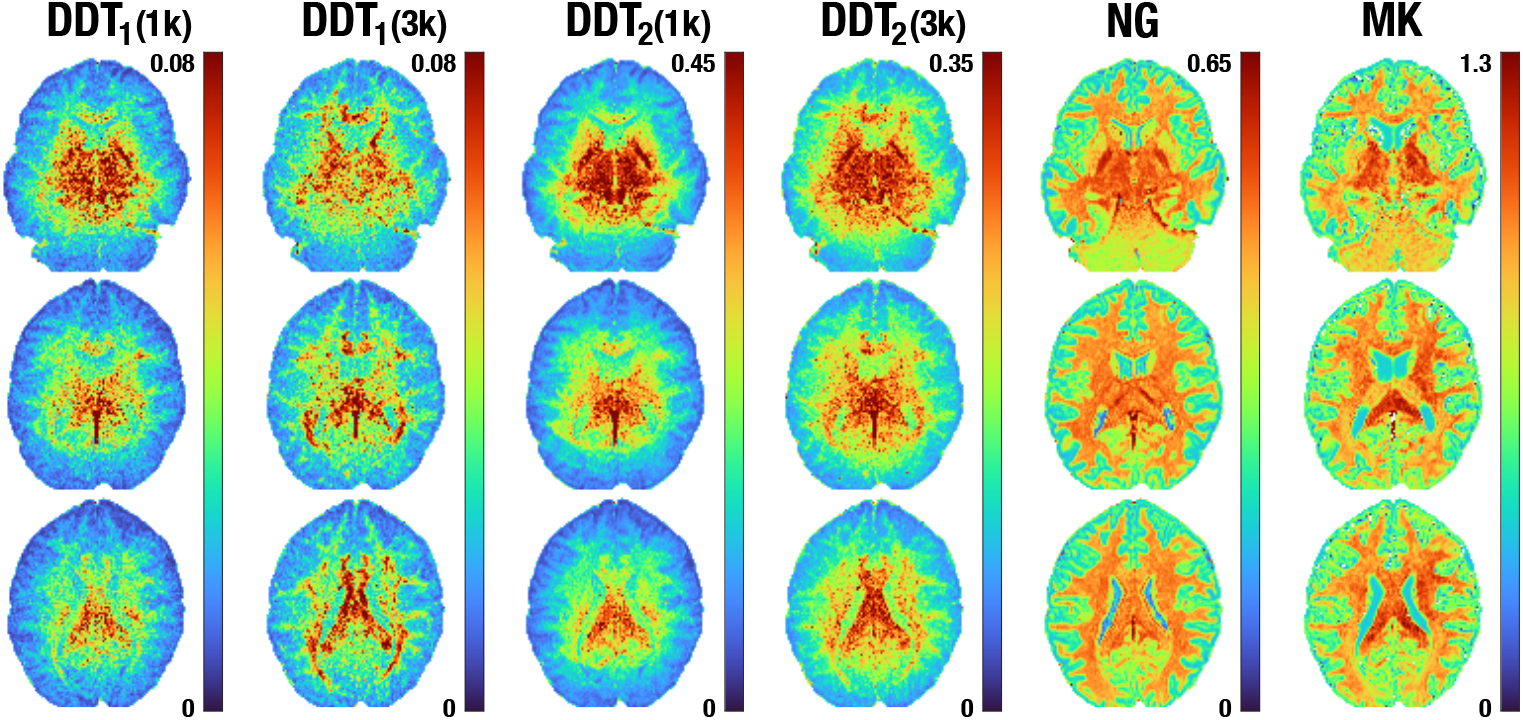
Quantification of different non-Gaussian diffusion metrics computed over the composite attenuation signal, subject HCP MGH1007. Each moment has been normalized to its own range for visualization purposes. DDT is calculated using a single shell at b=1000 s/mm^2^ and b=3000 s/mm^2^, NG using 3 shells (*b* = [1000, 3000, 5000] s/mm^2^) and MK using 2 shells (*b* = [1000, 3000] s/mm^2^)

Note that, although we use NG and MK as a reference, the three metrics do not describe the same effect. The first thing we can observe is that the levels of the three measures are quite different, even in the case of NG and DDT which, by definition, are in the interval [0,1]. We can, furthermore, observe that both NG and MK give a much clearer definition of the white matter. This is because the latter two metrics are based on multishell information, which allows them to distinguish where the signal does not follow a strictly Gaussian pattern. In the case of DDT, we are not measuring the same effect: we quantify how far the diffusion signal is from the best possible representation of that signal using a DT. When using a low shell, as in the case of *b* = 1000 s/mm^2^, many areas of the white matter are well represented by this tensor, so that a good part of it appears with low DDT values. It is precisely in those areas with a denser crossing of fibers in which the DDT values increase. These regions also correspond to the those with the highest values in NG and MK.

### 4.2 Synthetic Data Analysis

The main hypothesis of this work is that the proposed measure is able to detect differences in the white matter based on how the acquired signal diverges from the DT. In order to quantify this assumption, a synthetic experiment is carried out: we simulate two crossing fibers with an angle *α* inside a voxel. It is expected that the DT looses accuracy in modeling the diffusion process when the angle increases. For the sake of simplicity, we have considered three different scenarios, see Table 1 Synthetic Data Analysis for notation:

**TABLE 1.**
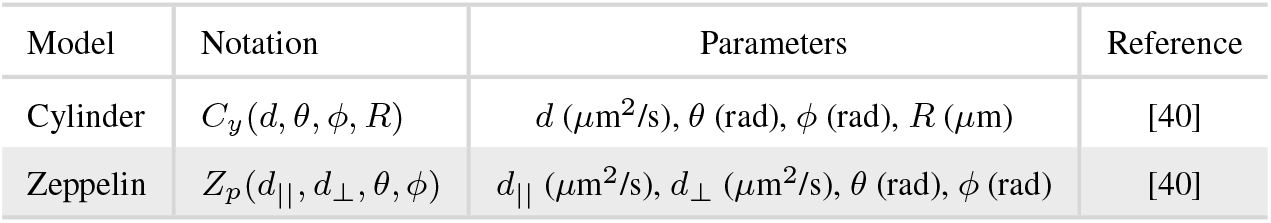
Compartment models used in the simulation.

1. The crossing of two zeppelin-shaped compartments:

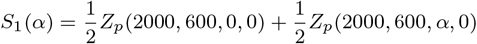
2. The crossing of three zeppelin-shaped compartments:

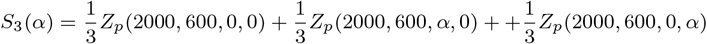
3. A more complex scenario, in which we assume that the intra-cellular compartment is modeled by a cylinder and the extra-celular is modeled by zeppelin. We consider a configuration with the crossing of 2 of such configurations:

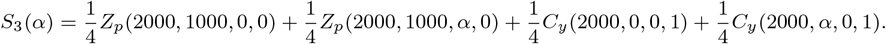

In order to simulate the diffusion signal, we consider the following sampling scheme: 61 gradient directions uniformly distributed in each shell, 1 baseline, 3 different shells at *b* = [1000, 2000, 3000] s/mm^2^, Δ = 35.5 ms, *δ* = 25.7 ms. For the sake of comparison we calculate DDT, NG and MK for different values of the crossing angle *α*. DDT is calculated using the lower shell (b=1000 s/mm^2^) while NG and MK are calculated using all the available information.

Results can be found in Fig 3 Synthetic Data Analysis. The first thing we notice in this case is that DDT exhibits much lower values than those of MK and NG. This effect was already apparent in Fig. 2 Visual Assessment, but when using synthetic data, it is amplified. Also note that by using larger values of b, larger output values of this parameter are also obtained.

**FIGURE 3.**
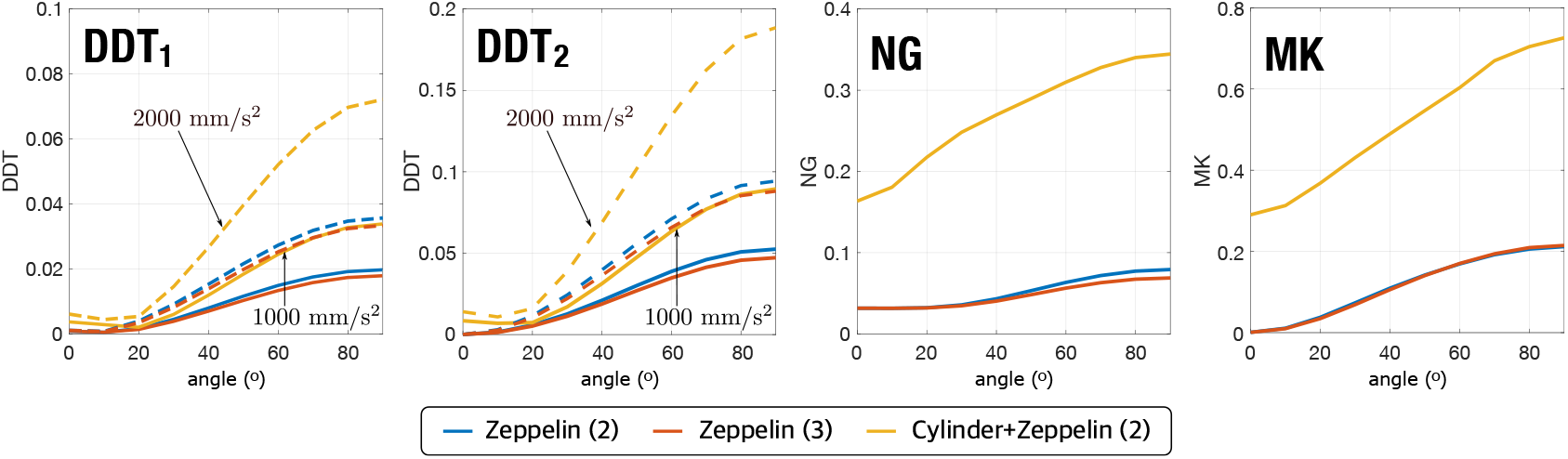
Results for the synthetic experiment with 2 crossing fibers at an angle *α*. Left to right: DDT_1_ calculated at b=1000 s/mm^2^ (continuous line) and b=2000 s/mm^2^ (dashed line); DDT_2_ calculated at b=1000 s/mm^2^ (continuous line) and b=2000 s/mm^2^ (dashed line); NG (calculated at *b* = [1000, 2000, 3000] s/mm^2^) and MK (calculated at *b* = [1000, 2000, 3000] s/mm^2^). Three different voxel configurations are considered.

In the case of the zeppelin models, when *α* = 0, the DT is able to perfectly model the diffusion within a voxel, so DDT has a null value. The same is true for MK while NG presents a small value, most likely due to numerical issues. When the angle increases, the value of DDT increases as well, since DT is no longer able to describe what happens inside the voxel. This growth also occurs, although more modest, in Mk and NG. Interestingly, MK provides the same values for the case of 2 and 3-fiber crossing, while DDT is able to distinguish between the two scenarios.

In the case of the zeppelin-cylinder scenario, even for zero angle, the diffusion cannot be correctly described by a diffusion tensor nor, in general, by a Gaussian. That is why all 3 methods give non-zero values in this case. When the crossing angle increases, all three measurements increases similarly. It should be noted that, in this case, DDT is able to distinguish the crossing using only information from a single shell. Both NG and MK distinguish the third case better than the previous one, since they have additional information provided by the use of several b values.

Following, we carry out a second synthetic experiment. We know that the higher order coefficient of SH are very sensitive to noise. In order to test how sensitive, we add some Rician noise to the synthetic signal *E*(**q**) before calculating the different metrics. First, we consider a simple zeppelin voxel with three different FA values, FA=[0.17, 0.41, 0.71], and different levels of noise, from 0% to 6%. For the sake of robustness, we consider the average and standard deviation of 1000 repetitions of each experiment. Results are on Fig. 4 Synthetic Data Analysis. We repeat the experiment for three different voxel configurations: 1 zeppelin, 2 crossing zeppelins and 2 cylinders and 2 zeppelin crossing.

**FIGURE 4.**
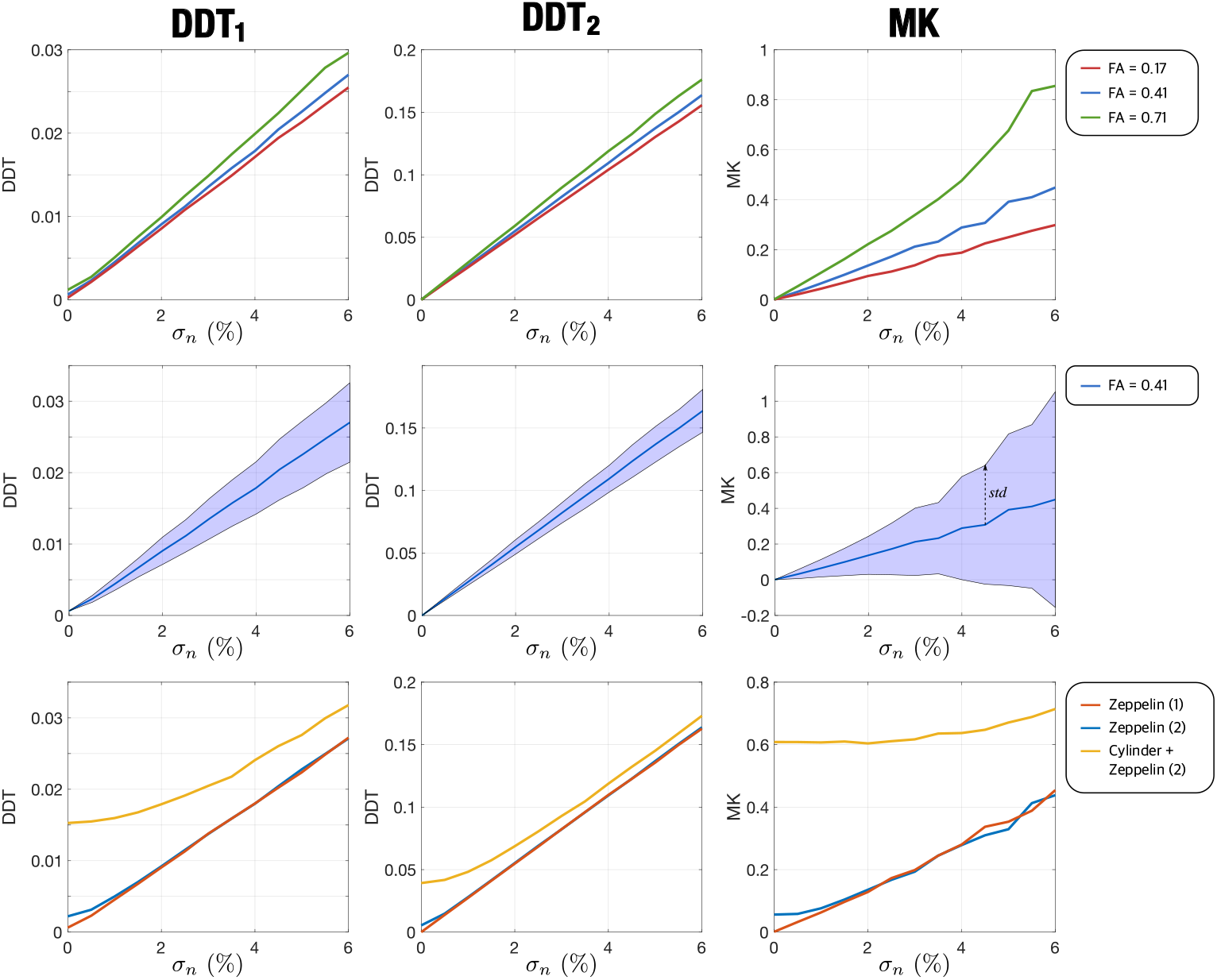
Results for the synthetic experiment with different levels of noise, *σ*_*n*_. Left to right: DDT_1_ calculated at b=1000 s/mm^2^; DDT_2_ calculated at b=1000 s/mm^2^ and MK calculated at *b* = [1000, 2000] s/mm^2^. Top row: zeppelin voxel with different FA (average). Middle row: zeppelin voxel with FA=0.41, average and standard deviation. Bottom row: Different voxel configurations (average). The results are obtained from 1000 repetitions of the same experiment.

First thing to notice is that the metrics are very sensitive to noise. The DDT shows a linear dependence with *σ*_*n*_, being DDT_2_ a bit more robust (in terms of smaller standard deviation). Noise can even mask the differences between the 3 voxel configurations. However, notice that a consolidate metric like the MK also shows a great noise dependence. However, its ability to resolve different voxels is higher, due to the usage of information from 2 shells.

### 4.3 Repeatability analysis

Having in mind the importance of a high level of reliability in neuroscientific research [42], we intend to make use of repeated-acquisitions data sets in order to complement the experiments carried out with a repeatability and separability assessment of DDT. To that end, we have used two publicly-available datasets: MICRA and HCP WU-Minn test-retest.

First, we assess the intra- and inter-subject variability of DDT using the MICRA data set and a region-oriented approach. DDT is calculated using only the shell at *b* = 1200 s/mm^2^. ROIs were back-projected by registering the JHU atlas’ labels [43] (in MNI152 space) into each subjects’ space by means of a linear and a nonlinear registration (FSL’s [44] FLIRT and FNIRT [45, 46], respectively) of the subject’s FA to the JHU atlas, followed by the application of the inverse warping of such to the JHU labels. Then, for each ROI in the subject space, the median of DDT values was computed, chosen as a single-valued representative of the measure enclosed within. Inter-subject variability was then computed as the variance across subjects of the means across sessions, while intra-subject was computed as the mean across subjects of the variances across sessions. Then, a figure of merit (FoM) consisting on the ratio between inter-subject and intra-subject variability was computed, i.e.

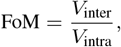

to get insights about the repeatability and reliability of the measures. For the sake of comparison, we have also calculated FA using the shell at *b* = 1200 s/mm^2^ and MK and NG using all the available shells.

Results can be seen in Table 2 Repeatability analysis (intra-subject CV).

**TABLE 2.**
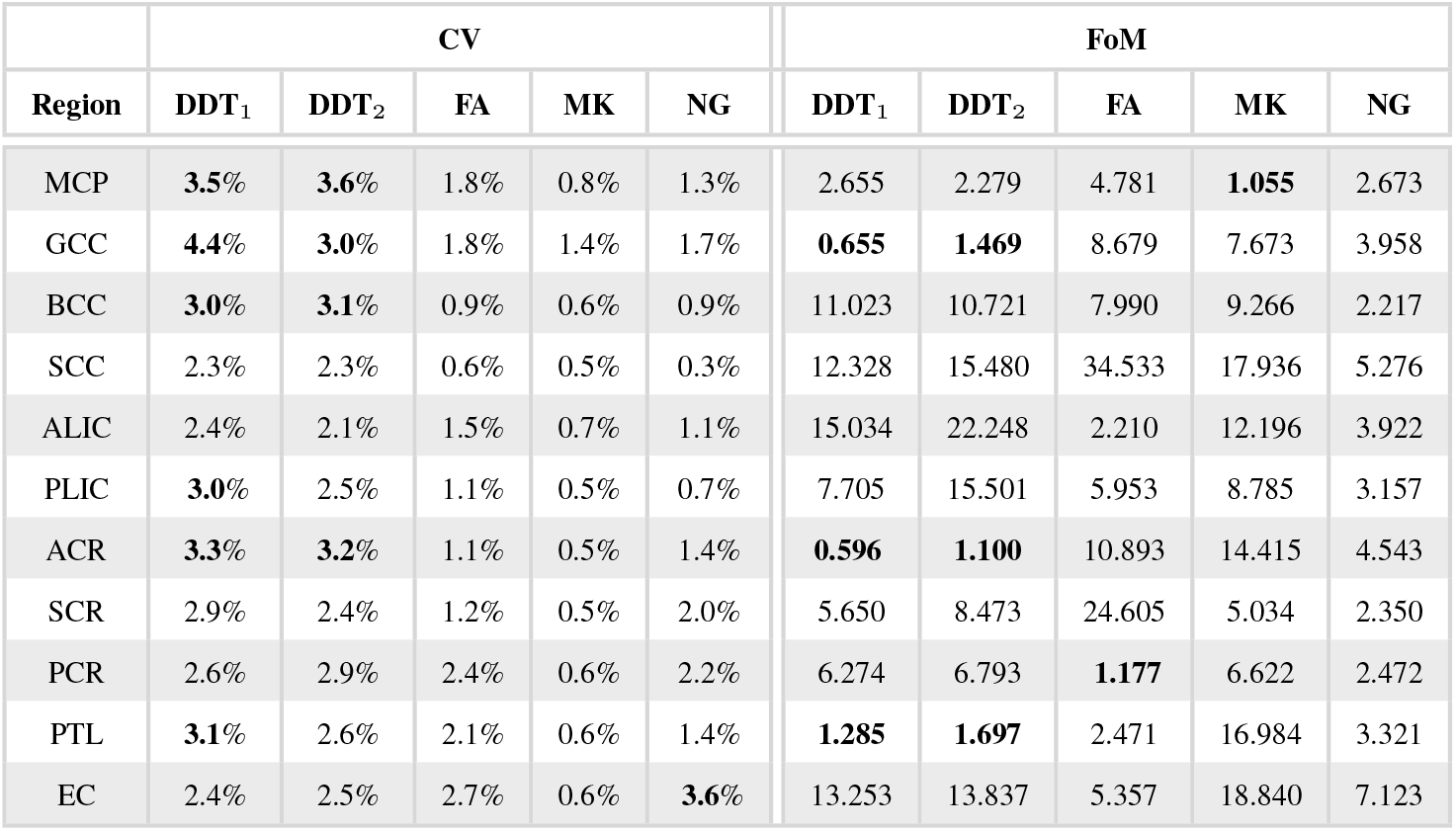
Repeatability results: Intra-subject Coefficient of Variation of DDT for the MICRA database (region oriented) and Figure of merit for each of the regions. Results indicate a good ratio between the inter-subject and the intra-subject variability for DDT. Lower values are highlighted.

Results shows a CV with a median value of 2.7% for the DDT, which is low enough to assure that the differences in the metric cannot be considered as noise. It is slightly higher than the CV of FA (also calculated using a single shell) and, as expected, higher than MK and NG, which use more information to be estimated. Nevertheless, the CV, although slightly higher, is in the same magnitude order of the other metrics.

Regarding the FoM, all the values are greater than one, meaning that, for any given ROI, the inter-subject variability of the metrics is greater than the intra-subject variability. It is also worth noticing that DDT outperforms NG in 2 of the 3 subregions of the *corpus callosum* (CC) —*Body* and *Splenium*— which together form one of the biggest connective pathways in the brain. Curiously, in the GCC DDT shows a low FoM when compared with all the other metrics.

Next, we repeat the study of the repeatability of DDT using the HCP test-retest database and a pointwise approach. To this end, we calculate the DDT under three configuration setups, i.e., *b* = 1000, 2000 and 3000 s/mm^2^, all using 90 gradient directions. We also obtain the FA parameter for each volume using FSL and non-linearly register them all to the FM-RIB58 template (a template averaged from 58 FA volumes provided by the FSL) using twelve degrees of freedom and a normalized correlation cost function [41]. Then, we warp the DDT measure calculated from all test and retest acquisitions to the standard space using trilinear interpolation. Once the measures are warped to the common space, we define the voxel-wise repeatability using the coefficient of variation (CV), that, for the specific case of 2 samples (test-retest) can be defined as

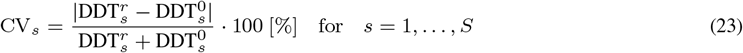

with CV_*s*_ being the voxel-wise CV calculated for subject *s* out of *S* (here, we assume *S* = 37), 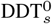 and 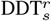 are respectively the test and retest acquisitions for each subject *s*. We obtain the global CV by aggregating the CVs retrieved from all subjects using a median operator: 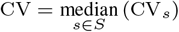.

Results of the experiment for a representative axial slice in the standard space are displayed in Fig. 5 Repeatability analysis. We observe a spatially-independent change of the CV of the DDT measure with the mean values at 5.18% (*b* = 1000 s/mm^2^), 4.71% (*b* = 2000 s/mm^2^) and 4.86% (*b* = 3000 s/mm^2^), all calculated for 20 representative slices of the brain.

**FIGURE 5.**
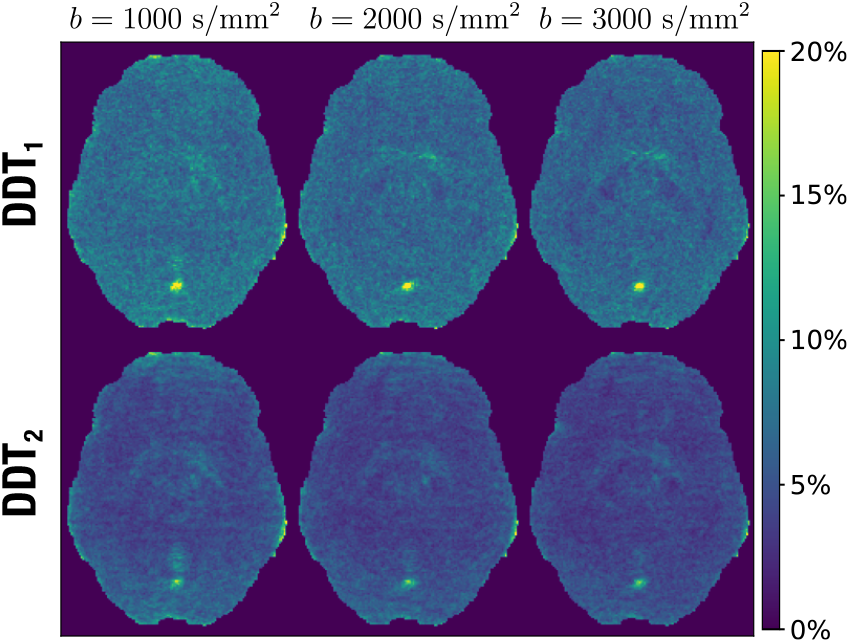
The CV maps of the DDT measures calculated under three different b-values, i.e., *b* = 1000, 2000 and 3000 s/mm^2^, all using 90 gradient directions. The CV maps were aggregated using a median operator from 37 subjects taken from the HCP WU-Minn test-retest database. The mean CVs calculated over 20 representative slices aggregated with a median operator over all subjects are (DDT_1_/DDT_2_): 7.48%/5.26% (*b* = 1000 s*/*mm^2^), 6.92%/4.69% (*b* = 2000 s/mm^2^) and 6.53%/4.35% (*b* = 3000 s/mm^2^).

## 5 DISCUSSION AND CONCLUSIONS

Although it is known that DT is not capable of correctly modeling what happens with diffusion within each voxel in a large part of the white matter, to date, many of the acquisitions performed in clinical studies are limited to a single shell in order to calculate the DT. Precisely in those regions where the DT fails, small variations in diffusion can be lost and, consequently, are not taken into account by the different clinical studies.

There are numerous techniques in the literature that allows the quantification of the non-Gaussian behavior of the diffusion in the white matter, such as those derived from kurtosis imaging. These techniques have shown that the quantification of how the diffusion diverges from the DT can be a significant indicator of various pathologies. However, the calculation of these metrics requires the use of additional b values.

In the present work we have proposed a new metric, DDT, which quantifies how the measured diffusion is separated from the diffusion reconstructed by the diffusion tensor, using only one b value.

While visually the measure does not resemble descriptors of non-Gaussianity, such as NG or MK, DDT provides higher values in those areas where the diffusion worst fits the tensor. We have seen that higher values of b refine the visual result. However, we believe that the measurement is more likely to be used at values of b around 1000 s/mm^2^, since it is then fully compatible with DTI.

In the synthetic experiment in Fig. 3 Synthetic Data Analysis, DDT has proved to be able to distinguish configurations with fiber crossings, as well as more complex scenarios with multi-chamber diffusion. Despite losing some information with respect to multishell approaches, it is able to provide extra information to that given by DT.

The repeatability of the measurement has been tested using two different databases, showing intra-subject variabilities of less than 5%, which evidences that the measurement is sufficiently robust. Furthermore, the FoM-based study shows that the between-subject variability is higher than the within-subject variability, which is key to be able to use it in future clinical studies. In addition, the visual result shows the independence of the CV from the brain region. This enables us to further translate the measure to the clinical applications or employ it in brain explorations across longitudinal acquisitions where high reproducibility is mandatory.

In summary, a new measure called DDT has been presented, a robust numerical formulation based on SH has been proposed and its performance has been validated.

## Data Availability

All data produced in the present study are available upon reasonable request to the authors.

## Software

The full implementation of DDT is included in the dMRImatlab toolbox and it may be downloaded for Matlab^©^ and Octave, together with use-case examples and test data, from: https://github.com/atriveg/dmrimatlab.

## Acknowledgments

This work was supported by Ministerio de Ciencia e Innovación PID2021-122528OB-100. Further support was received from MCIN/AEI/10.13039/ 501100011033 through grants PID2021-124407NB-I00, PID2024-162453NB-I00, and PID2024-158963NB-I00, co-funded by the European Union-European Regional Development Fund (ERDF, 2021?2027); and by the Junta de Castilla y León (grant VA156P24), co-financed by ERDF.Data collection and sharing for this project was provided by the *Human Connectome Project* (HCP; Principal Investigators: Bruce Rosen, M.D., Ph.D., Arthur W. Toga, Ph.D., Van J. Weeden, MD). HCP funding was provided by the National Institute of Dental and Craniofacial Research (NIDCR), the National Institute of Mental Health (NIMH), and the National Institute of Neurological Disorders and Stroke (NINDS). HCP data are disseminated by the Laboratory of Neuro Imaging at the University of Southern California.

## Conflict of interest

The authors declare that there is no conflict of interest.

## Footnotes

1 Data obtained from the Human Connectome Project (HCP) database (ida.loni.usc.edu/login.jsp). The HCP project (Principal Investigators: Bruce Rosen, M.D., Ph.D., Martinos Center at Massachusetts General Hospital; Arthur W. Toga, Ph.D., University of Southern California, Van J. Weeden, MD, Martinos Center at Massachusetts General Hospital) is supported by the National Institute of Dental and Craniofacial Research (NIDCR), the National Institute of Mental Health (NIMH) and the National Institute of Neurological Disorders and Stroke (NINDS). HCP is the result of efforts of co-investigators from the University of Southern California, Martinos Center for Biomedical Imaging at Massachusetts General Hospital (MGH), Washington University, and the University of Minnesota.

## Notes

### Competing Interest Statement

The authors have declared no competing interest.

### Author Declarations

The study used ONLY openly available human data that were originally located at: 1. Human Connectome Project (HCP) database from ida.loni.usc.edu 2. HCP WU Minn test retest database: from Van Essen DC et al. The WU Minn human connectome project: an overview. Neuroimage 2013 3. MICRA (Microstructural Image Compilation with Repeated acquisitions) from Koller K, Rudrapatna U, Chamberland M, Raven EP, Parker GD, Tax CMW, et al. MICRA: Microstructural image compilation with repeated acquisitions. NeuroImage 2021 jan

